# Atypical hyperendemicity of Rift Valley fever in Southwestern Uganda associated with the rapidly evolving lineage C viruses

**DOI:** 10.1101/2025.01.14.25320317

**Authors:** Barnabas Bakamutumaho, John Juma, Erin Clancey, Luke Nyakarahuka, Silvia Situma, Raymond Odinoh, Jeanette Dawa, Carolyne Nasimiyu, Evan A. Eskew, Stephen Balinandi, Sophia Mulei, John Kayiwa, John D. Klena, Trevor R. Shoemaker, Shannon L.M. Whitmer, Joel M. Montgomery, John Schieffelin, Julius Lutwama, Allan Muruta, Henry Kyobe Bosa, Scott L. Nuismer, Samuel O. Oyola, Robert F. Breiman, M. Kariuki Njenga

## Abstract

**Introduction:** Recent Rift Valley fever (RVF) epidemiology in eastern Africa region is characterized by widening geographic range and increasing frequency of small disease clusters. Here we conducted studies in southwestern (SW) Uganda region that has since 2016 reported increasing RVF activities.

**Methods:** A 22-month long hospital-based study in three districts of SW Uganda targeting patients with acute febrile illness (AFI) or unexplained bleeding was followed by a cross-sectional population-based human-animal survey. We then estimated RVFV force of infection (FOI) and yearly cases using the age-structured seroprevalence data and conducted genomic phylodynamic modelling of RVFV isolates.

**Results:** Overall RVF prevalence was 10.5% (205 of 1,968) among febrile or hemorrhagic cases, including 5% with acute (PCR or IgM positive) infection, averaging 5 cases per month. Community-based serosurvey recorded prevalence of 11.8% (88 of 743) among humans and 14.6% (347 of 2,383) in livestock. Expected yearly human RVF cases were 314-2,111 per 1,369 km*^2^* in SW Uganda versus 0-711 in comparable regions of Kenya and Tanzania. Viral genomic studies identified RVFV lineage C, sub-clade C.2.2, as the circulating strain in SW Uganda since 2019. Lineage C strain has undergone recent rapid evolution and clonal expansion resulting in four sub-clades, C.1.1, C.1.2, C.2.1, and C.2.2, that are more adept at establishing endemicity in new territories.

**Conclusions:** We demonstrate an atypical RVF hyperendemic region in SW Uganda characterized by sustained human clinical RVF cases, unusually high population prevalence, and high number of expected yearly human cases, associated in part with emergence of new RVFV sub-lineages.

**Key points:** Rift Valley fever (RVF) studies in SW Uganda found atypical sustained human cases averaging 5 cases/month, >10% population prevalence, and expected yearly cases >3-fold higher (314-2,111 vs 0-711) than comparable regions in East Africa, associated with emerging RVFV sub-lineages.

## INTRODUCTION

Rift Valley fever virus (RVFV) was first detected in Kenya in 1931, and subsequently across most countries in Africa over the next 70 years, resulting in severe human and livestock epidemics and serological evidence of endemicity in more than 30 countries [1–3]. In 2000, a severe RVF epidemic occurred in Saudi Arabia and Yemen in the Middle East, associated with livestock trade from the horn of Africa [4]. Once introduced to a country, this mosquito-borne virus becomes endemic in geographic areas supportive of virus maintenance; areas characterized by certain soil types, low elevation, and low annual rainfall [5–6]. In endemic countries, periodic epidemics, precipitated by heavy rainfall and flooding are followed by long (4-10 years) inter-epidemic periods (IEPs) with minimal disease activity [7–8].

Recent studies have highlighted two mechanisms associated with RVFV maintenance and disease burden during IEPs [9–12]. Apart from transovarial virus maintenance and transmission in mosquito eggs, RVFV is also maintained by continuous low-level circulation among wildlife, livestock, humans, and mosquitoes [10]. It is likely that dominance of this cryptic cycle is responsible for the changing epidemiology of RVF disease, which is characterized by a growing number of small RVF disease clusters (<5 human, <20 livestock cases) that occur across broader agro-ecological zones [13]. Unlike major regional epidemics, which often respond to El Niño Southern Oscillation (ENSO)-related extreme weather conditions, these small disease clusters during the IEP ebb and flow in response to routine seasonal climatic variation [11–14].

In the eastern Africa region, Kenya and Tanzania documented RVF endemicity decades ago and have been subjected to multi-country epidemics that also involved Somalia and Sudan, most recently in the epidemics of 1997-1998 and 2006-2007 [6, 15]. However, Uganda was not affected by these epidemics, reporting no human RVF cases even as the global and regional public health agencies enhanced regional surveillance during these epidemics [16]. The situation in Uganda changed in 2016 following confirmation of two RVF cases in the southwestern Kabale district [12]. Subsequent investigations detected anti-RVFV IgG antibodies in 13% of humans and livestock and infectious virus was detected in mosquitoes collected from the district [12]. Between 2016 and 2023, the country had reported >40 small human RVF clusters primarily in the southwestern and central districts of the country [11,13,17,18].

Here, we characterized the epidemiology of RVF disease in southwestern (SW) Uganda by conducting both a longitudinal hospital-based human study targeting patients with acute febrile illness or unexplained bleeding, and a cross-sectional population-based human-animal survey.

## METHODS

### Study area and design

We conducted two studies in the SW Uganda districts of Isingiro, Kabale, and Rubanda: a hospital-based human prospective study targeting patients ≥10 years old with acute febrile illness (AFI) and/or unexplained bleeding, and a cross-sectional community-based human-animal study. The 22-month (September 2021-July 2023) study enrolled febrile patients at Kabale Regional Referral Hospital in Kabale District, Hamurwa Health Centre in Rubanda District, and Rwekubo Health Centre in Isingiro District. Patients were enrolled if they exhibited undifferentiated fever (> 37.5*°* C) or a history of fever within the last 4 weeks and a negative malaria test. Additional criteria included unexplained bleeding or severe illness persisting for >7 days despite treatment. To investigate co-infection, 20% of the malaria positive participants were also enrolled. Our target sample size was 707 participants per site assuming a human 8% RVFV prevalence, with 80% power, 2% precision, and a 95% confidence level [12].

The cross-sectional survey was conducted in October-November 2023, using a two-stage randomization approach. First, 10 sub-counties were randomly selected across the three districts, and the number of households sampled within each sub-county allocated based on population density [19]. Subsequently, households were selected using randomly generated global positioning system (GPS) coordinates. At the household, one individual ≥ 1 year was randomly selected, interviewed, and blood sample collected. For a selected child (<10 years), a parent or guardian was interviewed. For households with livestock, a maximum of four animals of each species; (goats, cattle, and sheep) were randomly sampled. The target sample sizes, calculated using estimated RVF seroprevalence of 13% in humans, 17.7% in cattle, 3.7% in sheep, and 4.5% in goats, and factoring a 10% household non-response rate, was 982 humans and 2,476 livestock.

### Data and sample collection

After consenting, participants were clinically examined and both clinical, demographic, and risk factor data electronically collected in Research Electronic Data Capture (REDCap) software. Approximately 5 milliliters of whole blood were collected from each participant. Harvested sera and blood clots were aliquoted and shipped to Uganda Virus Research Institute (UVRI) and stored at −80°C for subsequent serological and molecular testing. Follow-up visits and sampling were carried out 4 – 6 weeks later.

### RVFV nucleic acid detection

Viral RNA was detected from serum using a previously published protocol [20]. Briefly, RNA was extracted using the MagMAX magnetic bead system (Life Technologies, Carlsbad, CA, USA) and amplification conducted using the following primer and probe set: 51’-TGAAAATTCCTGAGACACATGG-31’ (RVFL-2912fwdGG), 51’-ACTTCCTTGCATCATCTGATG-31’ (RVFL-2981revAC), and FAM-51’-CAATGTAAGGGGCCTGTGTGGACTTGTG-31’-BHQ (RVFL-probe-2950), on an ABI Quant Studio 5 real-time PCR platform (Thermofisher Scientific, Carlsbad, CA, USA).

### Anti-RVFV antibody detection

Sera were screened for total anti-RVFV antibodies (IgM and IgG) using a previously described protocol [11]. Briefly, heat and detergent inactivated sera were screened for RVFV total antibodies using a competition multi-species test designed to detect antibodies directed against the RVFV nucleoprotein (NP) in serum and plasma. The titers and the cumulative sum of optical densities for each dilution (SUMOD) minus the background absorbance of uninfected control antigen (adjusted SUMOD) were recorded. Samples were considered positive if the titer was 1:400 or above and the adjusted SUMOD and titer exceeded pre-established conservative cutoff values of ≥ 0.45 for IgM ELISA and ≥ 0.95 for IgG ELISA.

### Estimating RVFV force of infection in SW Uganda

To compare the burden of RVFV infection in SW Uganda with adjacent regions, we compared age stratified RVF population prevalence from our study (excluding hospital study) with four similar previously conducted in Kenya and Tanzania [13, 21–22]. Each study collected data on individual age, RVFV serological status, and geographic location. Individuals were assigned to 20 arcminute grid cells (roughly 37 km x 37 km in our study region) that encompassed our study region. Seventy-five grid cells containing serological data from at least 20 individuals we used as cutoff for inclusion in our analyses. These gridded serosurvey data were used to perform three analyses; (i)force of infection (FOI) for each grid cell using the age-structured serosurvey data, (ii) fit two gamma density functions to the FOI estimates, one for grid cells within the study area, (iii) used FOI estimates to calculate expected number of yearly RVF cases in each grid cell.

#### Estimating FOI

We estimated FOI for each grid cell using maximum likelihood, assuming (i) the FOI was constant over time and across age classes, (ii) disease-specific mortality was negligible, and (iii) that seroreversion was negligible [21, 23]. With these assumptions, the proportion of the population in each age class, *a*, that was seropositive was described by the following function:

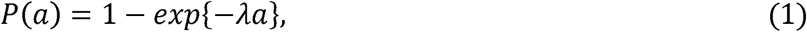

where *a* was measured in years and *λ* is the FOI. The likelihood of observing *x_i_* seroprositive individuals with corresponding age class *i* and 1 - *x_i_* seronegative individuals in age class *i* across age classes was:

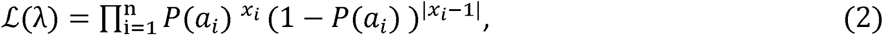

where *x_i_* is the serostatus {0,1} of individual *i* and *a_i_* is the age class, in years, of the *i*th individual. We maximized the negative logarithm of equation (2) with respect to *λ* in R using a variant of simulated annealing within the built-in function ‘optim’ [24,25].

#### Quantifying distribution of FOI

To more rigorously compare the distribution of estimated RVFV FOI experienced by human populations within SW Uganda to that experienced elsewhere in Tanzania and Kenya, we fit gamma density functions to the FOI data for each study region. Specifically, we used maximum likelihood to estimate the shape (*α*) and rate (*β*) parameters for gamma distributions fit to: 1) the FOI estimated for grid cells in Uganda, and 2) the FOI estimated for grid cells in Kenya and Tanzania. We again used simulated annealing in ‘optim’ and used the estimated shape and rate parameters to calculate the mode of the gamma distribution in each case as

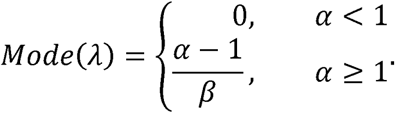

#### Estimating expected yearly RVF case counts

Expected RVF case counts were calculated by multiplying the FOI estimated for each grid cell by the estimated population size of seronegative humans in that cell. We generated human population size estimates by downloading population density data for the study region from the WorldPop dataset (https://hub.worldpop.org/) [26], converting density data to human population counts, and then aggregating the count data to the 20 arcminute grid cell size. We relied on WorldPop data from 2020, the most recent year available. All manipulation of raster data used functions from the *terra* R package [27].

### Whole genome sequencing and phylogenetic analysis of RVFV isolates

High throughput next-generation sequencing was performed on 5 of the 10 RVF PCR-positive samples with Ct ≤25 (samples CD300/Kabale1-2022, CD301/Rubanda1-2022, CD304/Rubanda2-2023, CD532/Isingiro1-2023) using a protocol described previously [28–30]. Briefly, RNA was extracted from blood or serum using 5X Magmax™ 96 Viral Isolation kit and libraries prepared following unbiased shotgun method. Samples were sequenced using RVFV-specific primers designed from Uganda-specific RVFV sequences and the ARTIC protocol [28–29]. Consensus genome sequences were constructed using the ARTIC bioinformatics protocol using RVFV-specific config files to a RVF reference genome sequence (GenBank accession # MG972978).

#### Genomic data retrieval, filtering, and lineage assignment

Publicly-available RVFV sequence data from the National Center for Biotechnology Information (NCBI) database (search limited to Uganda) were combined with data generated from this study [31]. We successfully retrieved genomic sequence data for the 3 fragments, excluding vaccine strains: S (n = 33), M (n = 24), and L (n = 30). For each segment sequence dataset, we determined RVFV lineages using the lineage assignment tool [32].

#### Phylogenetic analysis

Multiple sequence alignment of each dataset was performed using MAFFT [33] followed by manual editing using Aliview [34] for correct codon-alignment. We inferred the best substitution models using ModelTest-NG [35] and identified HKY+G4, GTR+I, GTR+G4 as optimal for the S, M, and L segments, respectively. We incorporated these evolutionary models in maximum likelihood phylogenetic tree inference using IQ-TREE [34] while estimating branch support values using ultrafast bootstrapping procedure (-bb 1000). To deduce the temporal signal of the sequence data, we regressed the genetic distances and the sampling dates (in years) through a root-to-tip plot (**Supplementary Figure 1**). We further utilized the sampling dates to compute time-scaled phylogenetic trees using TreeTime [36]. Our root-to-tip reconstructions indicated that the L segment data contained sufficient temporal signal (Coefficient correlation = 0.99 and R*^2^* = 0.99) to warrant Bayesian inference. We therefore used the L segment sequence data to perform phylogeographic analysis. To deduce the spatial diffusion of RVFV in Uganda, we utilized continuous phylogeographic reconstruction in BEAST [37], with incorporation of uncorrelated log-normal tree branching and skyline coalescent demographic models. Using *tidygeocoder* [38], we geocoded the geographical sampling locations into longitude and latitude and used the coordinates as traits in a Cauchy distribution model for continuous phylogeographic inference. BEAST was run with 100 million Markov chain Monte-Carlo (MCMC) steps, sampling every 10,000*^th^* step. Effective sample size values (ESSs > 200) were assessed using Tracer [39] to ensure proper convergence and mixing of the outputs. We used Tree Annotator [40] to retrieve and annotate the maximum clade credibility (MCC) tree. We extracted the spatial-temporal information embedded in these phylogeographically reconstructed trees using the *seraphim* package in R [39] to ascertain RVFV transmission in Uganda.

### Ethical and administrative approvals

Ethical approvals for this study were provided by the Uganda Virus Research Institute Research Ethics Committee (Study # GC/127/849) and the Uganda National Council for Science and Technology (Study # HS1713ES).

## RESULTS

### Sustained detection of acute RVF cases in SW Uganda

Overall, 2,711 participants were enrolled in both the hospital-based (n = 1,968) and cross-sectional community (n = 743) studies. Between October 2021 and July 2023 (22 months), a total of 1,968 febrile patients were enrolled at three health facilities located in Isingiro, Kabale, and Rubanda districts, out of which 100 (5.1%) were clinically positive by either viral RNA (n = 6), antiviral IgM antibodies (n = 90), or both (n = 4). Of these 100 clinically positive participants, 90 were managed as outpatients while 10 had severe disease requiring hospitalization. The major clinical manifestations included fever (92%), headache (84%), joint pains (64%), chills (61%), fatigue/weakness (53%), and bleeding (10%) (**Figure 1**). None of the participants died. Interestingly, there was sustained detection of clinical RVF cases throughout the 22-month period, averaging 5 (range 1-13) cases per month, but ranging between 0-20% of AFI patients weekly as illustrated in ***Figure 1***.

**Figure 1.**
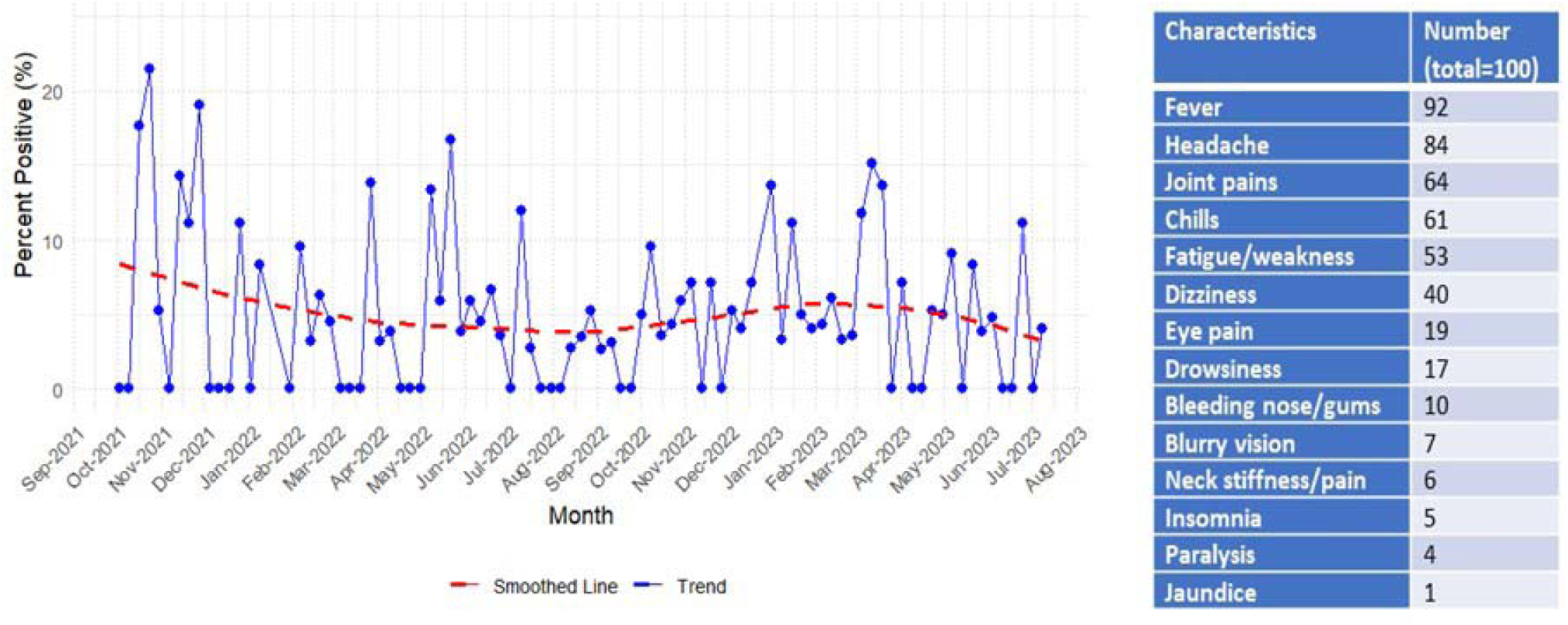
Epicurve (A) and clinical characteristics (B) of acute RVF cases detected at three health facilities in SW Uganda between September 2021 and July 2023 (n = 100).

No clinically positive RVF cases (viral RNA or IgM positive) were detected in the cross-sectional study (n = 743) conducted across the 3 districts.

### Human and livestock RVF seroprevalence

Of the 2,711 human participants tested for RVFV ribonucleic acid or IgM and IgG antibodies, 293 (10.8%) were positive, including 205 of 1,968 (10.4%) from the prospective hospital-based study and 88 of 743 (11.8%) from the cross-sectional study. We observed clustering of acute cases near the three hospitals where surveillance was conducted (**Figure 2A&B, red dots**), whereas the distribution IgG positive participants was more widespread across the 3 districts (**Figure 2B, green dots**). Of 2,383 livestock tested from the cross-sectional study, 347 (14.6%) were positive for anti-RVF IgG antibodies, including a significantly higher (p < 0.05) seroprevalence rate in cattle (33.8%, 230 of 681) than in goats (6.7%, 78 of 1170) or sheep (7.3%, 39 of 532).

**Figure 2.**
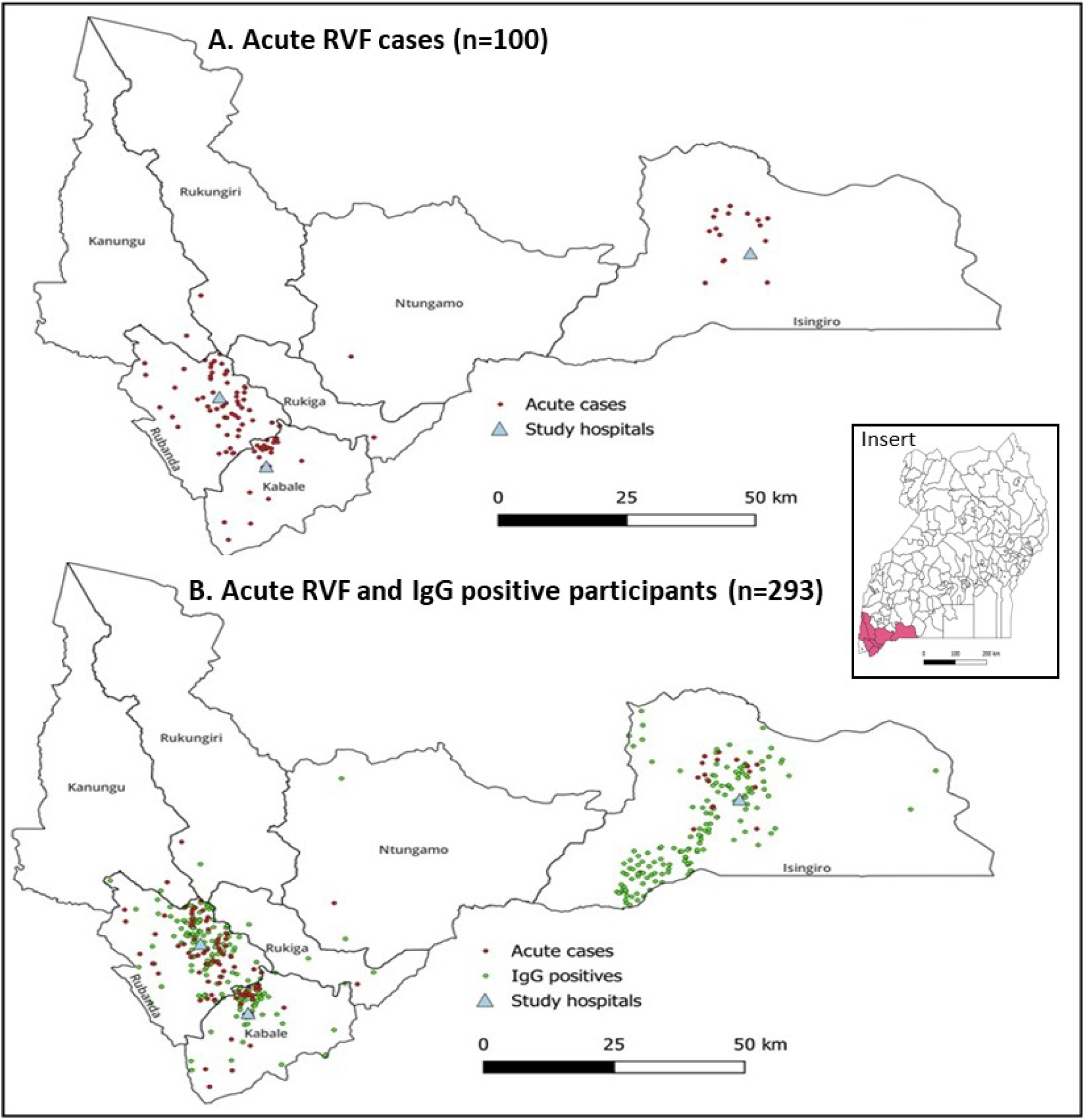
Spatial distribution of human acute RVF cases (A), and both acute RVF (red dots) and IgG positive (green dots) participants (B) in the Southwestern Uganda region. <u>Insert:</u> Map of Uganda showing the SW Uganda region.

### RVFV force of infection and expected yearly RVF cases

To determine the burden of RVFV infection in the region, we compared our estimates for the force of infection (FOI) acting on human populations at our study sites to estimates for adjacent regions in Kenya and Tanzania, using data compiled from previous serosurveys [13,21–22]. Although the range of FOI estimates was similar between the two regions, the distributions differed (**Figure 3A),** as did the expected yearly RVF cases **(Figure 3B).** Specifically, fitting gamma density functions to the FOI estimates revealed a modal value of 0.00096 in SW Uganda but 0 in Kenya and Tanzania (**Figure 3C**). This difference in the shape of the distributions shows that most human populations sampled in SW Uganda are exposed to RVFV whereas most human populations sampled outside this region are unexposed to RVFV. These results are consistent with widespread, sustained circulation of low-to-medium levels of RVF disease in SW Uganda but isolated and sporadic episodes of intense RVFV infection in Kenya and Tanzania.

**Figure 3.**
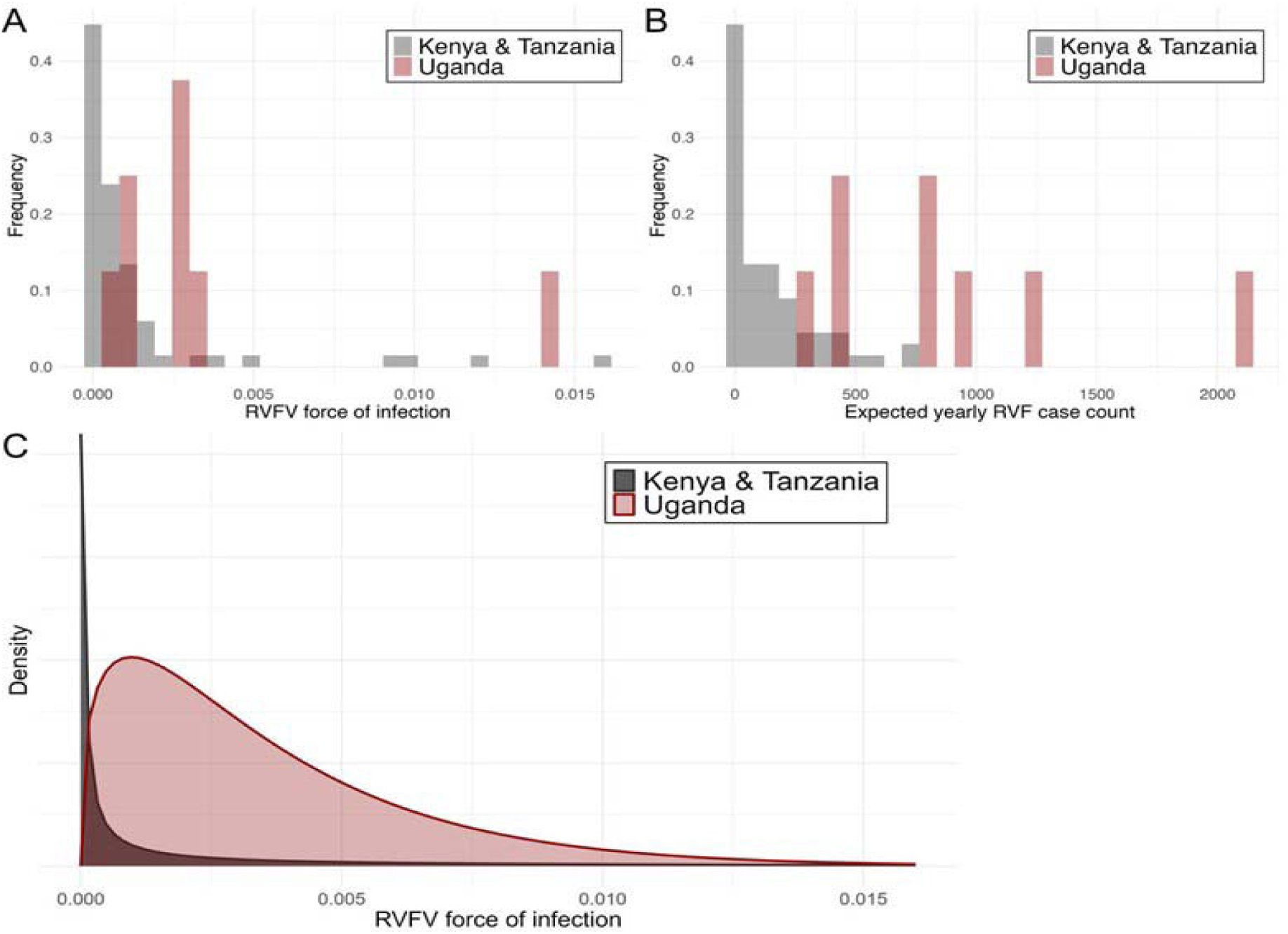
Frequency distribution of RVFV force of infection (A), expected yearly RVF cases per grid cell (B), and gamma density functions fit to the force of infection estimates (C). The gamma density functions fit to force of infection estimates from each region show that the characteristics of the distributions are different, driven by consistent virus circulation during IEP in in SW Uganda, and minimal circulation in adjacent regions of Kenya and Tanzania.

After transforming estimated FOI to expected RVF case counts in each grid cell, we estimated that the SW Uganda region could expect 314-2,111 yearly RVF cases per grid cell, whereas the Kenya and Tanzania region could expect 0-711 yearly RVF cases per grid cell (**Figure 4**).

**Figure 4.**
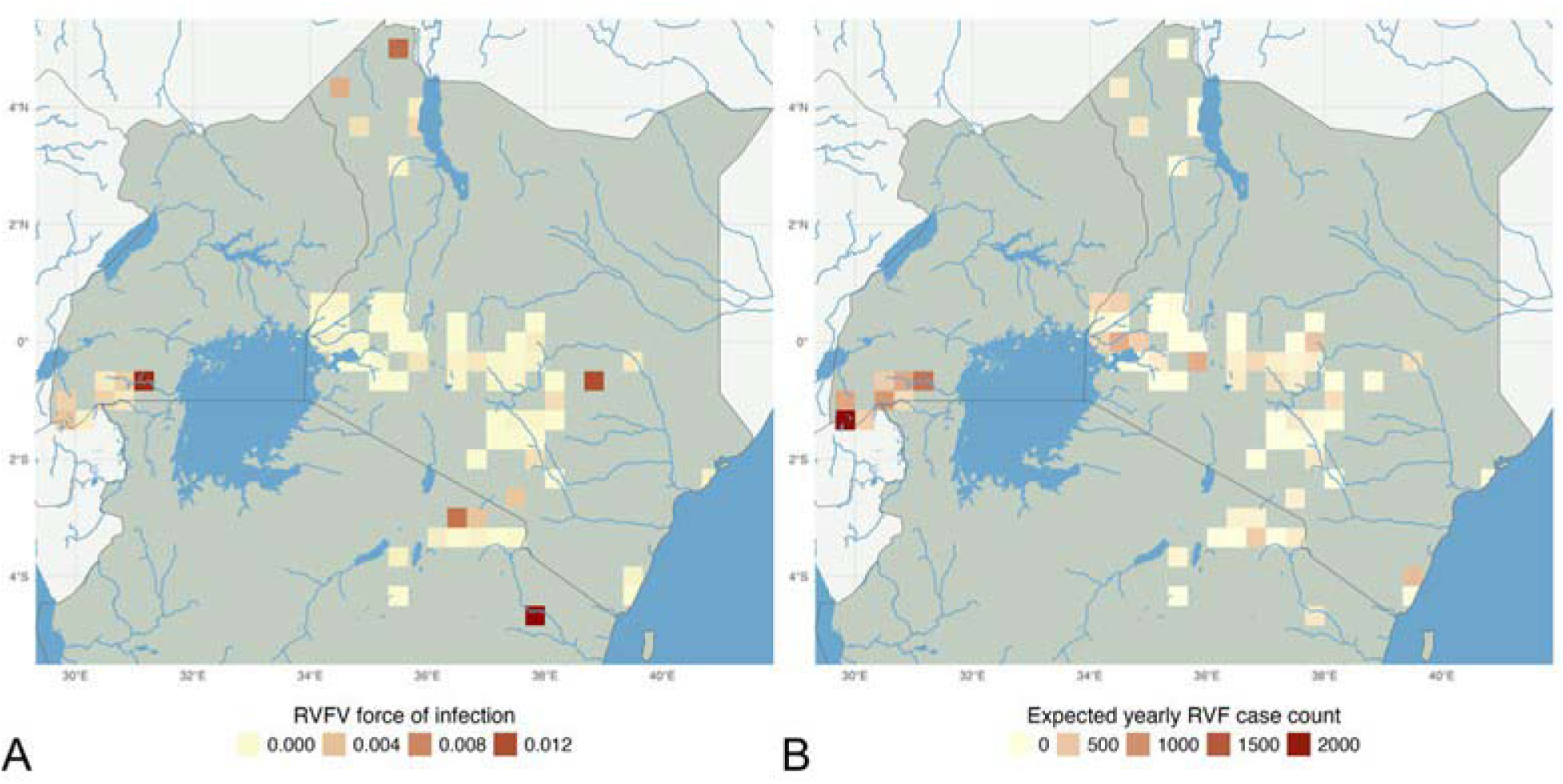
Estimated RVFV force of infection (FOI) for different spatial locations in Kenya, Uganda, and Tanzania (A) and translation into expected yearly RVF cases (B). The figure illustrates higher RVF circulation the southwest Uganda region, which has a high FOI and large human population, leading to high expected yearly RVF cases.

### Circulating RVFV lineages

From the 10 PCR positive samples, whole genome sequencing was successful for four samples: two from the Rubanda and one each from Isingiro districts and Kabale districts (**Supplementary Table 1**). The samples were collected between October 3, 2022, and January 30, 2023, from male participants, 26-30 years old. The S (non-structural genes), M and L segments of the isolates aligned with RVFV lineage C, sub-clade 2.2 (**Figure 5, Supplementary Tables 2,3,4**).

**Figure 5.**
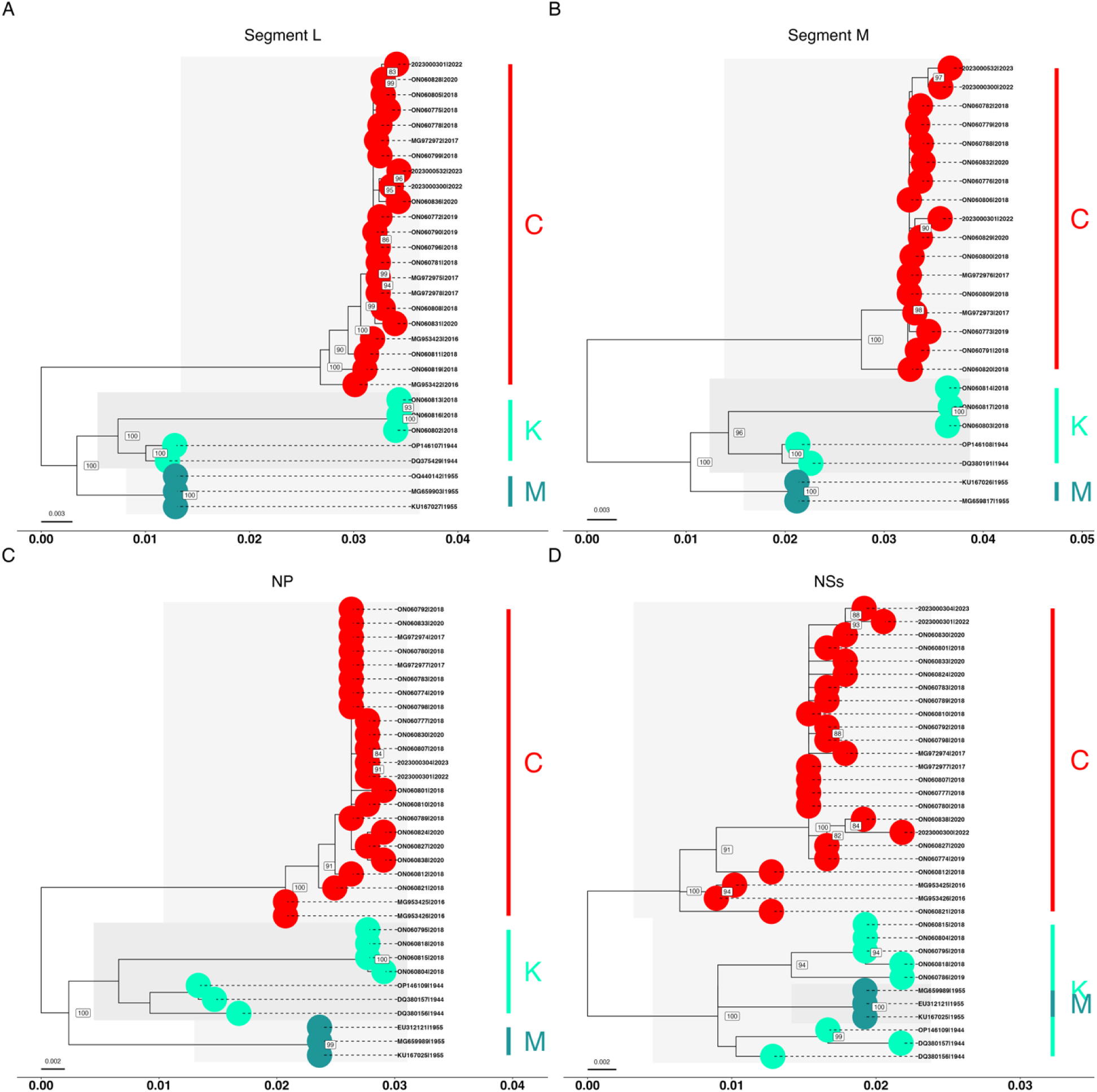
Phylogenetic inference of RVFV lineage evolution in Uganda. Maximum likelihood phylogenetic trees were reconstructed using IQ-TREE. (**A**) L segment (n = 30), (**B**) M segment (n = 24), (**C**) nucleoprotein (n = 33) and (**D**) non-structural gene sequences from the S segment. The tips of the trees are colored using the RVFV lineage information from assignment and labelled by accession numbers and sampling dates (year). Statistical support for tree nodes are indicated by bootstrap values.

#### Evolution of RVFV lineages in Uganda

We examined the evolutionary relationships of RVFV lineages circulating in Uganda (using genomic sequence data collected between 1944 and 2023). Maximum likelihood phylogenetic trees show that between the years 1940 and 2000 the circulating strains of RVFV were of lineages C, K (Entebbe) and M (Lunyo) (**Figure 5**). Phylogenetic inference shows clustering of lineages K and M followed by emergence of lineage C in several parts of the country from around 1990. Lineage C, shown to be predominant in Uganda since 2015 has undergone expansion into distinct sub-lineages C.2.1 and C.2.2 [51]. Notably, sub-lineage C.2.1 emerged in the country during the first reported human outbreak in 2016 followed by expansion to sub-lineage C.2.2. Interestingly, we observed long branches in a cluster of 3 sequences isolated in 2018 belonging to lineage K (here reported as K.1.2) sharing ancestry with the livestock vaccine Smithburn isolated in Entebbe in 1944 [**Figure 5**]. Although there is a huge gap in sampling RVFV between 1950s and 2000s (a potential bias in sampling) the temporal signals as examined by regressing root to tip genetic distances provide sufficient statistical support for the current Bayesian evolutionary analysis (**Supplementary Figure 1**).

#### Phylogeographic dispersal of RVFV lineages in Uganda

We established sufficient temporal signal in the large segment sequences and applied continuous phylogeographic analysis to understand the dispersal dynamics of RVFV lineages in Uganda. Although our genomic dataset was limited in terms of the number of sequences sampled (n = 30), the phylogeographic analysis show that RVFV lineages tend to disperse from SW Uganda region. The dispersal diffusion of RVFV lineages showed several introductions of the virus through the SW Kabale and Isingiro districts and eastern Busia district (**Figure 6**). We showed that the time of the most recent ancestor (tMRCA) of RVFV lineages in Uganda may have been over 100 years, 1921 [95% HPD: 1899 – 1940]. The evolution rate of the medium segment is 3.57E-4 [95% HPD: 2.62E-4 – 4.65E-4] substitutions per site per year (subs/site/year). Our continuous phylogeographic analysis aligns well with the analysis patterns of livestock movements in the so-called cattle corridor, which stretches from southwestern to northeastern Uganda.

**Figure 6.**
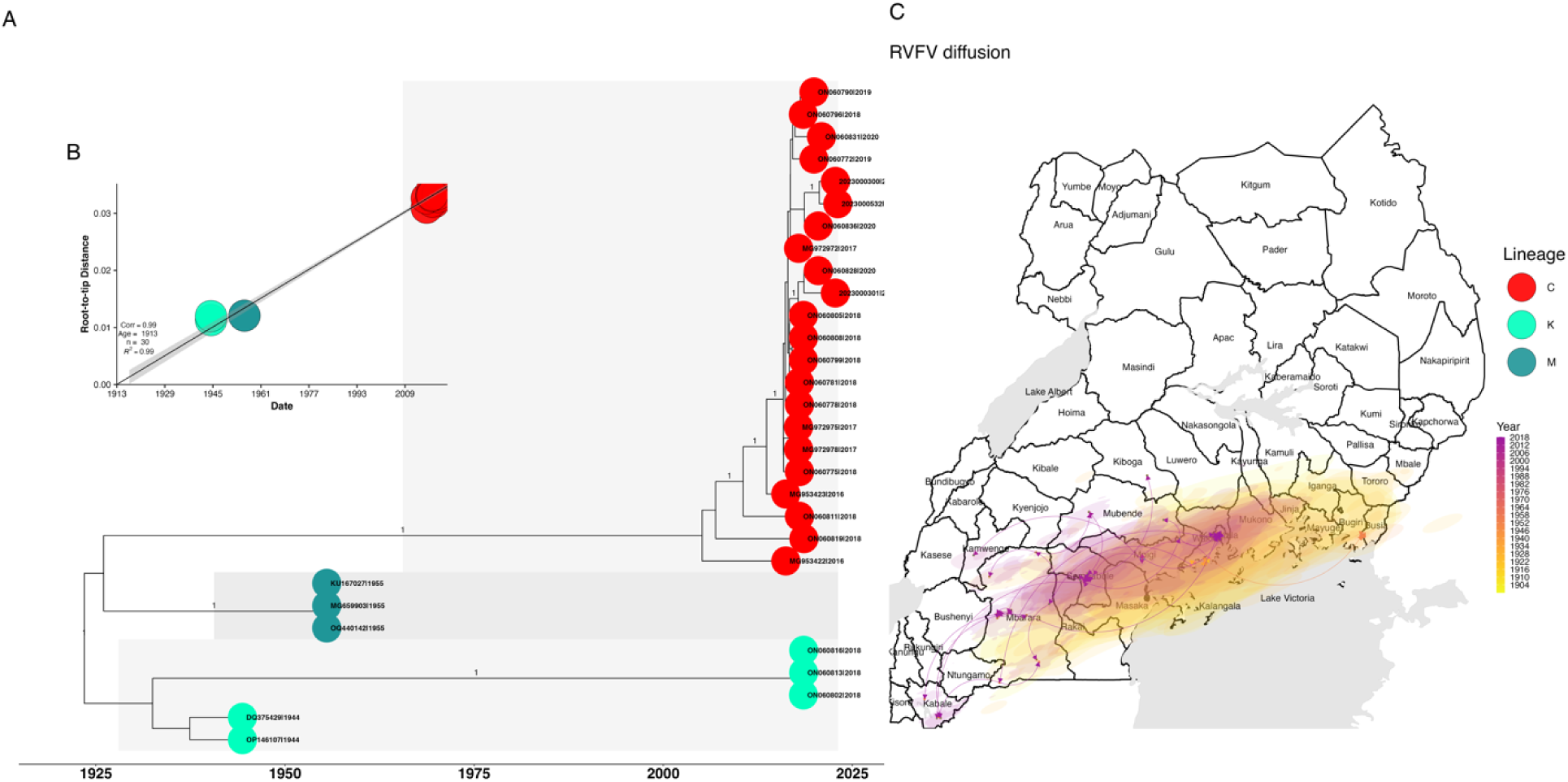
Continuous phylogeographic analysis of RVFV lineages in Uganda using complete L segment sequences (n = 30). Dispersal history of RVFV lineages summarized by a maximum clade credibility (MCC) tree retrieved and annotated from 900 posterior trees sampled from the posterior distribution of the continuous phylogeographic analysis (**A**) Maximum clade credibility (MCC) tree retrieved from the Bayesian phylogenetic inference based on the medium segment genomic sequences and highlighting the clustering of the different lineages of the virus. The tips of the tree are colored according to lineage information and labels show the accession numbers and sampling time in years. (**B**) Root-to-tip plot showing regression of the genetic distances and the sampling time (year) of the isolates. (**C**) Nodes of the trees are colored according to the time of occurrence, and oldest nodes are plotted on top of youngest ones. The light grey shaded areas in the map depict lakes.

## DISCUSSION

To our knowledge, this is the first study describing an RVF hyperendemic region characterized by sustained detection of human clinical RVF cases streaming to hospitals, unusually high population prevalence among humans and livestock, and estimated FOI consistent with high expected annual human RVF case counts across the region. Over a period of almost 2 years, 5% (100 of 1,968) of patients presenting with acute febrile illness or unexplained bleeding at three hospitals in SW Uganda were positive for RVF, averaging 4.5 cases per month and up to 20% of febrile cases per week. While none of the patients died, they exhibited classic RVF clinical manifestations, including headache, joint pains, and bleeding syndromes, with 10% requiring hospitalization for intensive clinical care. An additional 5% (105 of 1,968) of the enrolled patients had prior exposure to RVFV, demonstrated by presence of antiviral IgG antibodies, giving an overall RVF disease prevalence of 10.5% (205 of 1,968) among febrile patients in the region. As expected, our spatial mapping showed clustering of these cases around the hospitals involved in the study, suggesting RVFV infection is likely to be widespread in the region given the challenges associated with seeking health care in rural Uganda.

Within the community, we found human and livestock RVFV seroprevalences of 11.8% (88 of 743) and 14.6% (347 of 2,383), respectively. Our similar studies in the Central highlands of Kenya and the eastern Democratic Republic of Congo failed to detect acute RVF cases among febrile patients, while in communities human and livestock RVFV prevalences were 3-6-fold lower than in SW Uganda [41]. When compared to the few randomized population-based seroprevalence studies conducted elsewhere, the RVFV prevalence in SW Uganda was 2-6-fold higher [21,41–42]. Comparing the distribution of RVFV FOI estimates between human populations in SW Uganda with adjacent regions in Kenya and Tanzania showed an interior mode of 0.00096 in SW Uganda while it was 0 in other regions, indicating that SW Uganda had persistently non-zero FOI, while most areas in Kenya and Tanzania had a zero or near zero FOIs. Overall, these findings confirm existence of sustained low-to-medium levels of RVF cases in SW Uganda.

Virus genomic evolution and transmission studies showed that the RVFV strains circulating in SW Uganda since 2019, including isolates from the current study, belongs to lineage C sub-lineage C.2.2, which was first detected in Kenya during the 2006/2007 RVF epidemic [32, 43]. Previous studies showed that lineage C is the predominant virus circulating in eastern Africa region since 1990s [44]. First detected in Zimbabwe in 1976, lineage C virus was introduced to Kenya thereafter undergoing clonal expansion during the large 2006-2007 epidemic and creating distinct sub-clusters. Phylogenetic analysis focusing on lineage C indicates a variant undergoing evolution at a relatively increased rate, producing four sub-lineages, C1.1, C1.2, C2.1, and C2.2. This relatively rapid evolution could be attributed to silent circulation during IEPs. Phylogeographic studies shows the ability of lineage C to establish endemicity in new territories, likely therefore catalyzing the widening geographic range of small RVF clusters recently described [13].

Although lineage K was not detected among the current isolates, our evolutionary analysis shows that there was occurrence of a new lineage K cluster (here referred to as K.1.2) in Uganda in 2018 that is related to the livestock vaccine strain, Smithburn virus. The occurrence of a vaccine-related strain in human population may indicate a possible expansion of this lineage. Continuous phylogeographic inferences place southeastern and southwestern Uganda as the centers of origin of RVFV transmission to other parts of the country. The occurrence of these sustained RVF disease activities at border points with other countries such as Rwanda, Kenya and DRC require enhanced surveillance especially at these entry points.

Apart from virus strains, other factors that likely contribute to RVF hyperendemicity in SW Uganda include climatic changes and anthropogenic factors such as land-use-change and environmental degradation. Our recent study correlated long-term climatic trends; rising annual mean temperature (at 0.12*°* to 0.3*°*C per decade) in East Africa and increasing rainfall trends in scattered highlands of the regions (especially annual number of rainy days), including southwestern highlands of Uganda with ocurrent RVF clusters [13].

## Supplementary Data

Supplementary materials are available at *Clinical Infectious Diseases* online. Consisting of data provided by the authors to benefit the reader, the posted materials are not copyedited and are the sole responsibility of the authors, so questions or comments should be addressed to the corresponding author.

## Notes

## Acknowledgements

We thank the Uganda Ministry of Health for administrative approval and the Uganda Virus Research Institute (UVRI) for laboratory support and ethical oversight. We thank Alex Tumusiime, Jackson Kyondo, and Justine Okello for field and diagnostic support, and Angela Ndiu and Stella Nabatanzi for administrative support.

## Authors’ Contributions

B. B., L. N., J. D., R. F. B., and M. K. N. conceived and designed the study. B. B., J. J., E. C., L. N., S. S., R. O., J. D., C. N., E. A. E., S. B., S. M., J. K., J. D. K., T. R. S., S. L. M. W., J. M. M., J. S., J. L., A. M., H. K. B., S. L. N., S. O. O., R. F. B., M. K. N.; drafting and revision of the manuscript. All authors reviewed and approved the final version of the report. All authors accept responsibility to submit for publication.

## Financial support

Funding for this project was provided by the US National Institute of Allergy and Infectious Disease/National Institutes of Health (NIAID/NIH), grants number U01AI151799 through the Centre for Research in Emerging Infectious Diseases-East and Central Africa (CREID-ECA). Genomic work in Dr Samuel Oyola’s laboratory was supported by the Global Health EDCTP3 (Grant Agreement no. 101103171) Joint Undertaking and its members as well as Bill & Melinda Gates Foundation and the Rockefeller Foundation. Laboratory testing at UVRI was supported by US Centers for Disease Control and Prevention through a grant to the institute. Silvia Situma received training support from the NIH/Fogarty International Center’s D43 training grant # D43TW011519 awarded to Washington State University and University of Nairobi. John Schieffelin was supported by NIAID/NIH grant # 5U01AI151812 through the West Africa Research Network in Infectious Diseases (WARN-ID).

## Data availability

All decoded data generated or analyzed in this study will be available upon reasonable request. GenBank accession numbers for sequences derived in this study were PQ436077 through PQ436088.

## Potential conflicts of interest

All authors report no conflicts of interest.

## Supplementary data

**Supplementary Table 1.**
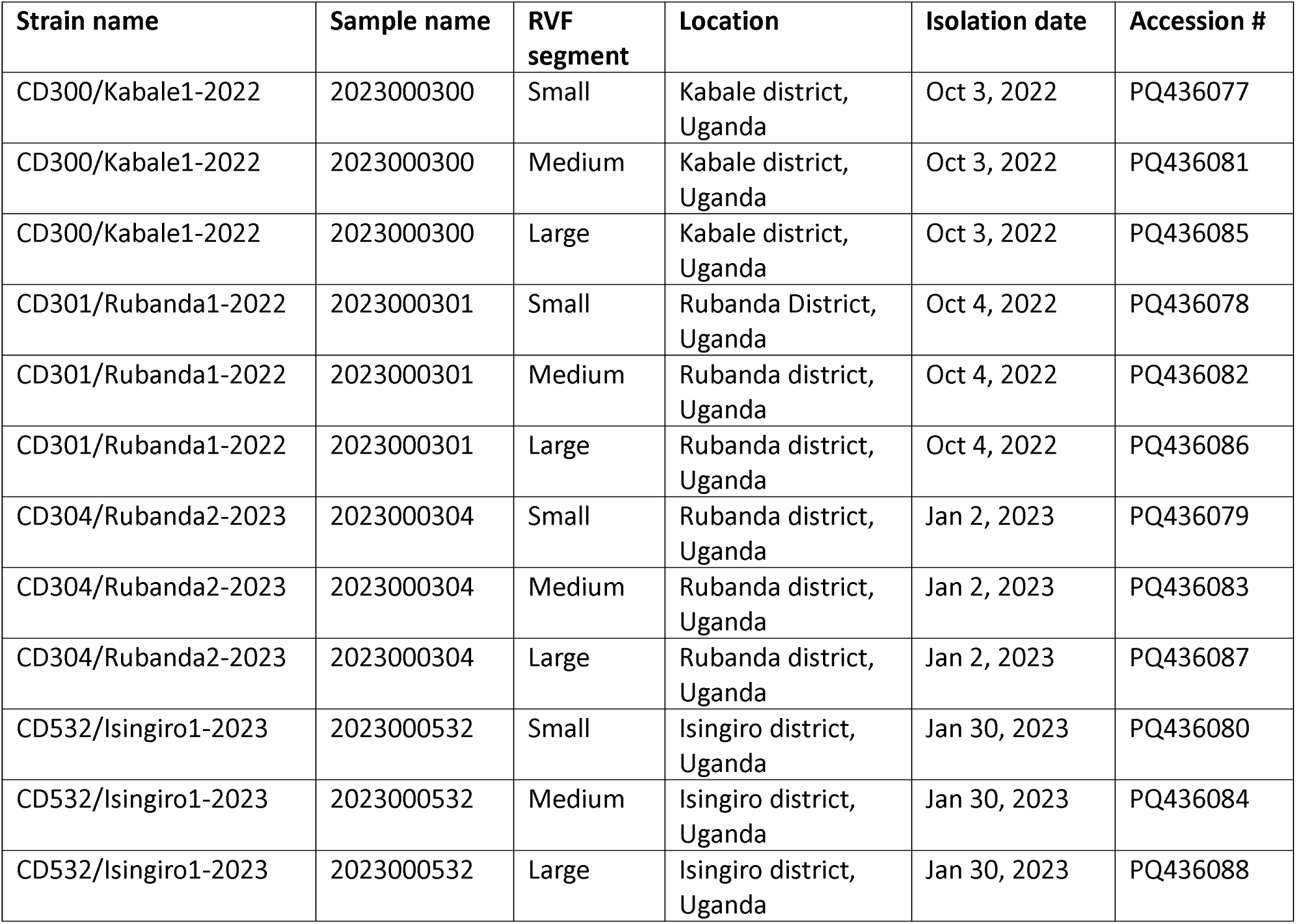
Names and GenBank accession number of the four RVFV strains isolated in southwestern Uganda, 2022-2023.

**Supplementary Figure 1.**
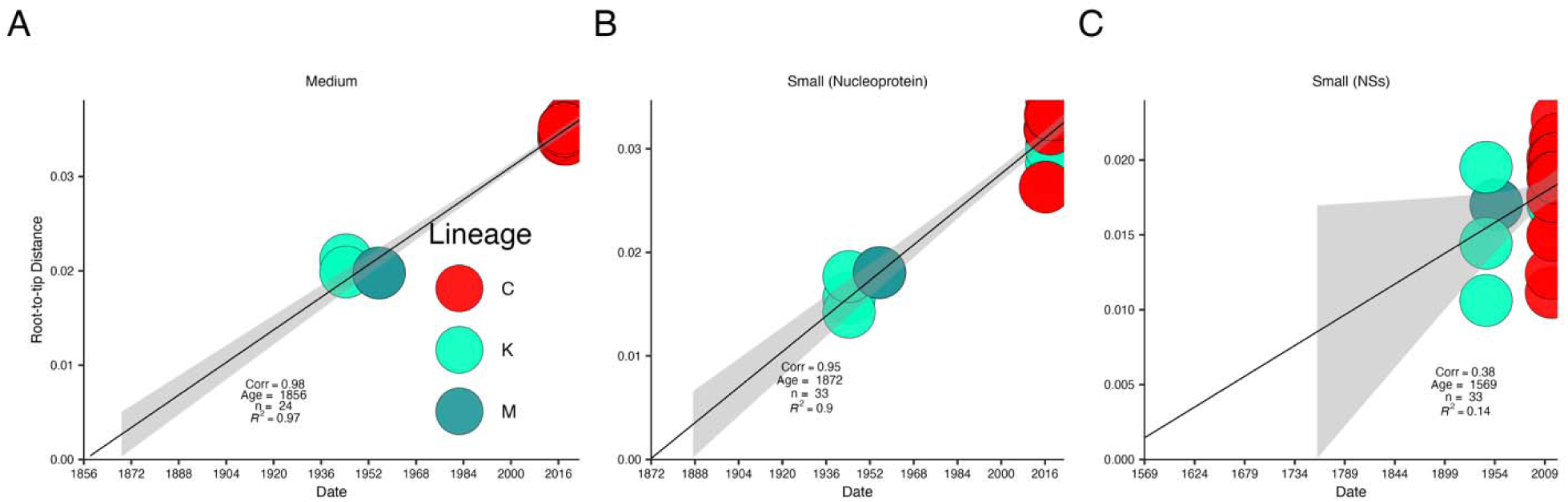
*Temporal signal assessing the root to tip distance versus sampling time in years.* Pairwise genetic distances were computed between the oldest sequence (root) and each sequence in the dataset. (**A**) RVFV regression of the root-to-tip genetic distances versus the sample collection dates using complete (**A**) medium genomic sequences and (**B**) nucleoprotein and (C) non-structural gene sequences in the small segment.

**Supplementary Table 2.**
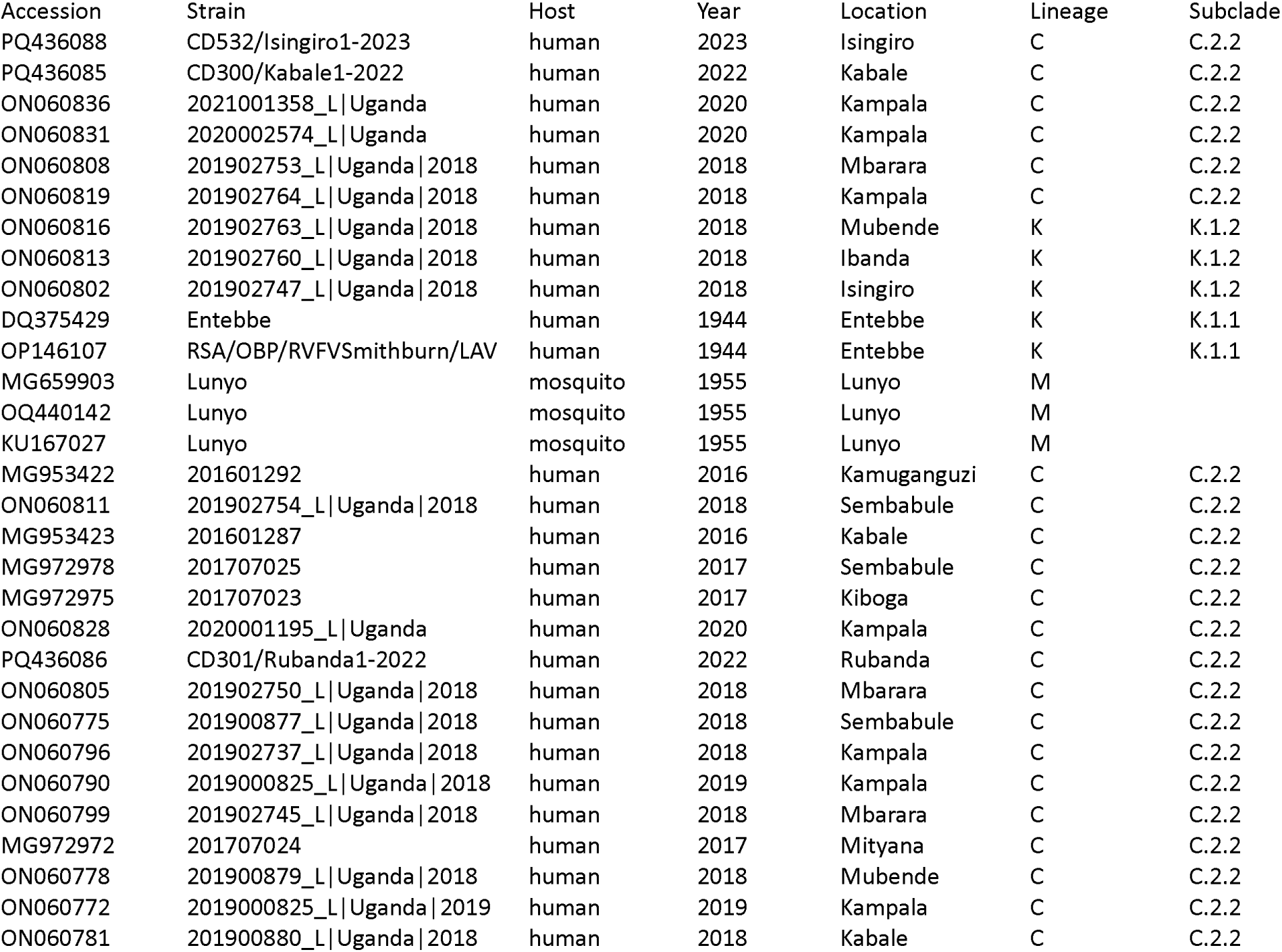
Lineage assignment of complete large segment genomic sequences.

**Supplementary Table 3.**
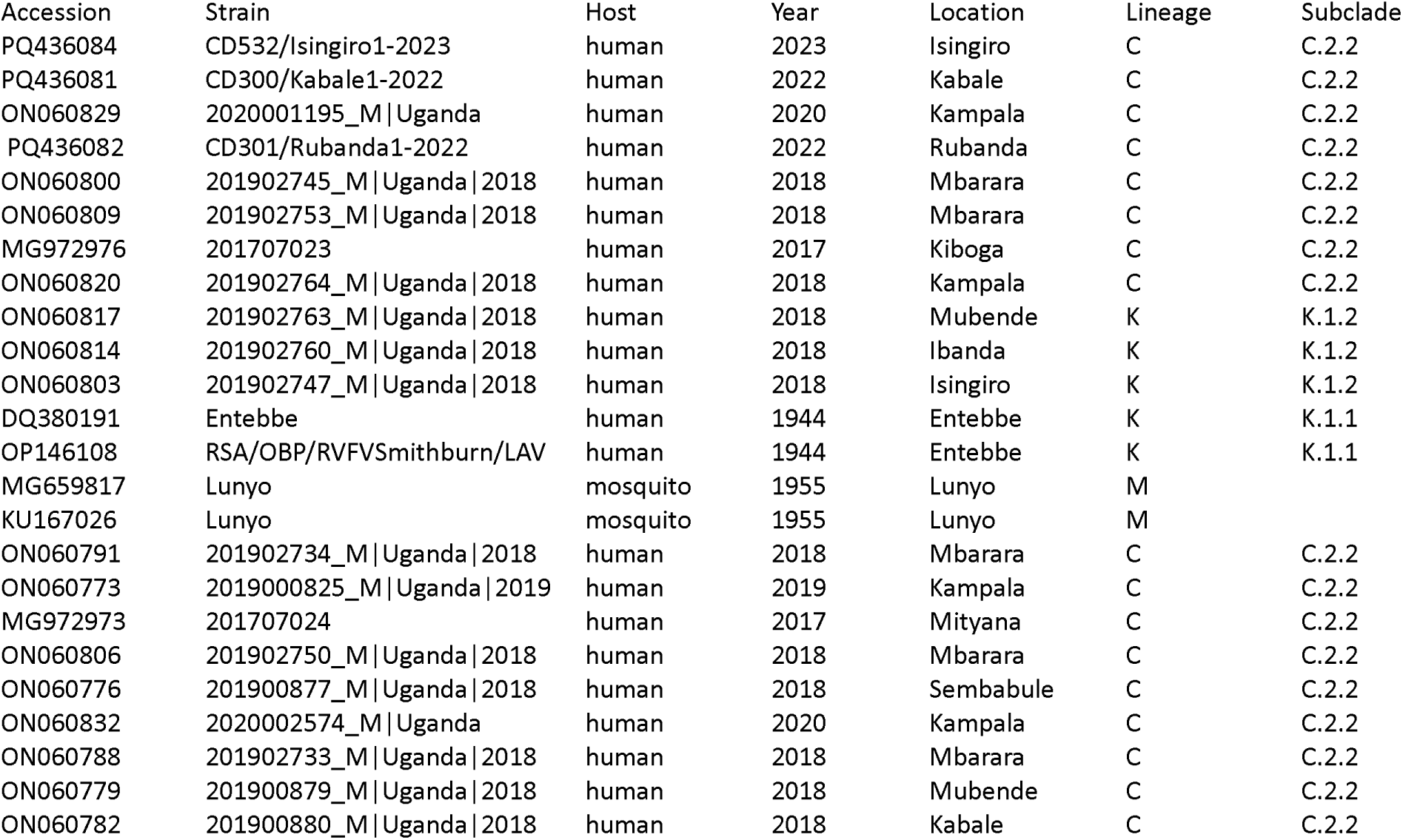
Lineage assignment of complete medium segment genomic sequences.

**Supplementary Table 4.**
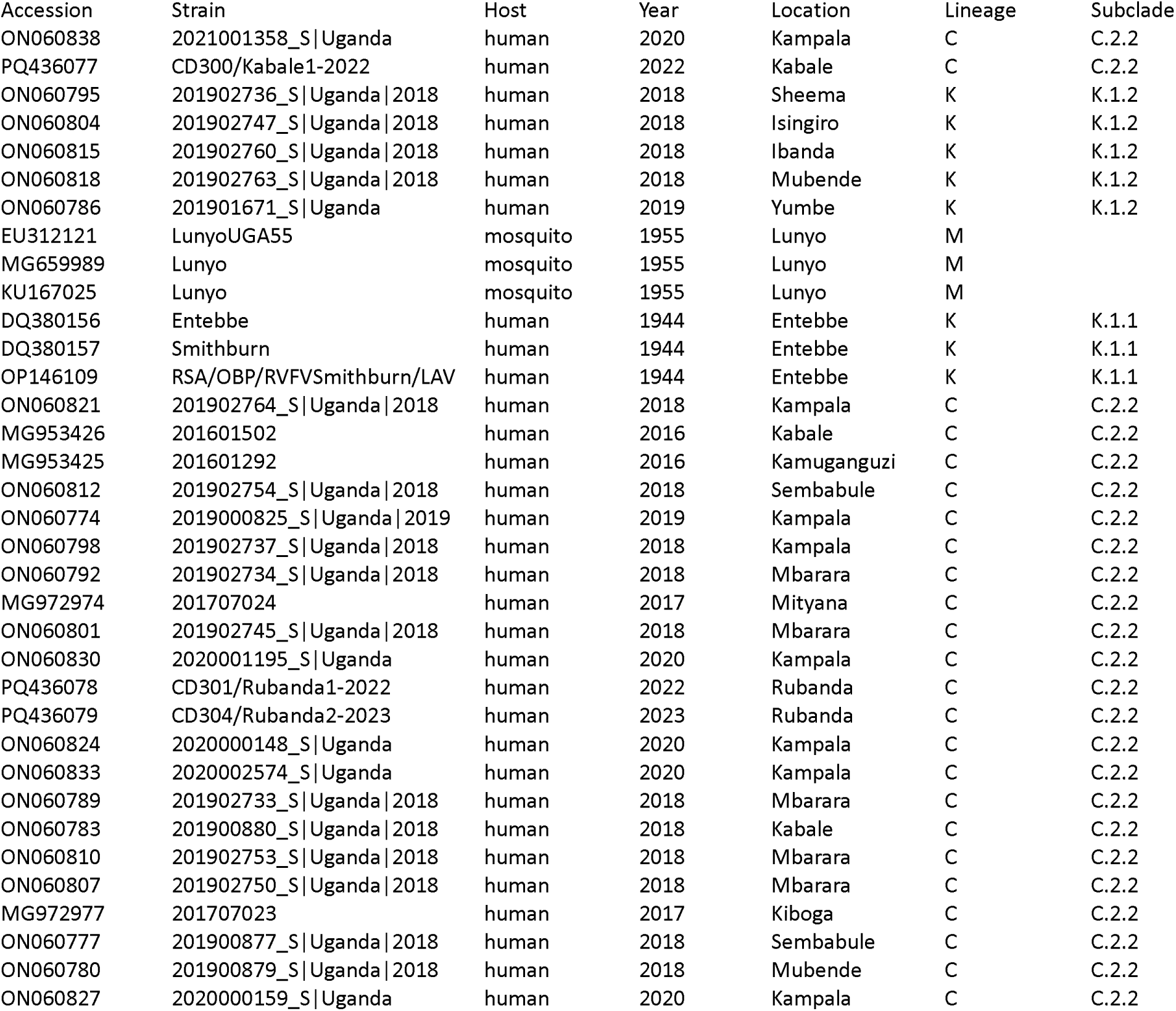
Lineage assignment of complete small segment genomic sequences.

## Notes

### Competing Interest Statement

The authors have declared no competing interest.

### Author Declarations

Ethical approvals for this study were provided by the Uganda Virus Research Institute Research Ethics Committee (Study Number GC/127/849) and the Uganda National Council for Science and Technology (Study Number HS1713ES).

