## Supplemental data for "Atypical hyperendemicity of Rift Valley fever in Southwestern Uganda associated with the rapidly evolving lineage C viruses"

**Supplementary data**

**Supplementary Table 1.** Names and GenBank accession number of the four RVFV strains isolated in southwestern Uganda, 2022-2023.

| **Strain name** | **Sample name** | **RVF segment** | **Location** | **Isolation date** | **Accession #** |
| --- | --- | --- | --- | --- | --- |
| CD300/Kabale1-2022 | 2023000300 | Small | Kabale district, Uganda | Oct 3, 2022 | PQ436077 |
| CD300/Kabale1-2022 | 2023000300 | Medium | Kabale district, Uganda | Oct 3, 2022 | PQ436081 |
| CD300/Kabale1-2022 | 2023000300 | Large | Kabale district, Uganda | Oct 3, 2022 | PQ436085 |
| CD301/Rubanda1-2022 | 2023000301 | Small | Rubanda District, Uganda | Oct 4, 2022 | PQ436078 |
| CD301/Rubanda1-2022 | 2023000301 | Medium | Rubanda district, Uganda | Oct 4, 2022 | PQ436082 |
| CD301/Rubanda1-2022 | 2023000301 | Large | Rubanda district, Uganda | Oct 4, 2022 | PQ436086 |
| CD304/Rubanda2-2023 | 2023000304 | Small | Rubanda district, Uganda | Jan 2, 2023 | PQ436079 |
| CD304/Rubanda2-2023 | 2023000304 | Medium | Rubanda district, Uganda | Jan 2, 2023 | PQ436083 |
| CD304/Rubanda2-2023 | 2023000304 | Large | Rubanda district, Uganda | Jan 2, 2023 | PQ436087 |
| CD532/Isingiro1-2023 | 2023000532 | Small | Isingiro district, Uganda | Jan 30, 2023 | PQ436080 |
| CD532/Isingiro1-2023 | 2023000532 | Medium | Isingiro district, Uganda | Jan 30, 2023 | PQ436084 |
| CD532/Isingiro1-2023 | 2023000532 | Large | Isingiro district, Uganda | Jan 30, 2023 | PQ436088 |

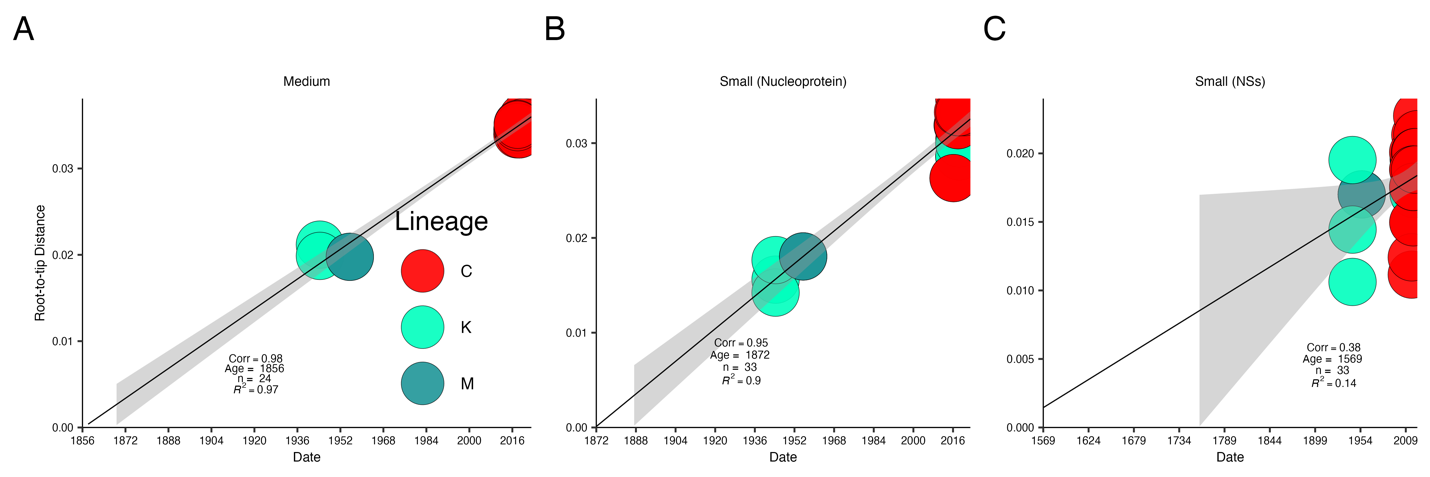

***Supplementary Figure 1****. Temporal signal assessing the root to tip distance versus sampling time in years.* Pairwise genetic distances were computed between the oldest sequence (root) and each sequence in the dataset. (**A**) RVFV regression of the root-to-tip genetic distances versus the sample collection dates using complete (**A**) medium genomic sequences and (**B**) nucleoprotein and (C) non-structural gene sequences in the small segment.

**Supplementary Table 2**. Lineage assignment of complete large segment genomic sequences.

| Accession | Strain | Host | Year | Location | Lineage | Subclade |
| --- | --- | --- | --- | --- | --- | --- |
| PQ436088 | CD532/Isingiro1-2023 | human | 2023 | Isingiro | C | C.2.2 |
| PQ436085 | CD300/Kabale1-2022 | human | 2022 | Kabale | C | C.2.2 |
| ON060836 | 2021001358_L\|Uganda | human | 2020 | Kampala | C | C.2.2 |
| ON060831 | 2020002574_L\|Uganda | human | 2020 | Kampala | C | C.2.2 |
| ON060808 | 201902753_L\|Uganda\|2018 | human | 2018 | Mbarara | C | C.2.2 |
| ON060819 | 201902764_L\|Uganda\|2018 | human | 2018 | Kampala | C | C.2.2 |
| ON060816 | 201902763_L\|Uganda\|2018 | human | 2018 | Mubende | K | K.1.2 |
| ON060813 | 201902760_L\|Uganda\|2018 | human | 2018 | Ibanda | K | K.1.2 |
| ON060802 | 201902747_L\|Uganda\|2018 | human | 2018 | Isingiro | K | K.1.2 |
| DQ375429 | Entebbe | human | 1944 | Entebbe | K | K.1.1 |
| OP146107 | RSA/OBP/RVFVSmithburn/LAV | human | 1944 | Entebbe | K | K.1.1 |
| MG659903 | Lunyo | mosquito | 1955 | Lunyo | M |  |
| OQ440142 | Lunyo | mosquito | 1955 | Lunyo | M |  |
| KU167027 | Lunyo | mosquito | 1955 | Lunyo | M |  |
| MG953422 | 201601292 | human | 2016 | Kamuganguzi | C | C.2.2 |
| ON060811 | 201902754_L\|Uganda\|2018 | human | 2018 | Sembabule | C | C.2.2 |
| MG953423 | 201601287 | human | 2016 | Kabale | C | C.2.2 |
| MG972978 | 201707025 | human | 2017 | Sembabule | C | C.2.2 |
| MG972975 | 201707023 | human | 2017 | Kiboga | C | C.2.2 |
| ON060828 | 2020001195_L\|Uganda | human | 2020 | Kampala | C | C.2.2 |
| PQ436086 | CD301/Rubanda1-2022 | human | 2022 | Rubanda | C | C.2.2 |
| ON060805 | 201902750_L\|Uganda\|2018 | human | 2018 | Mbarara | C | C.2.2 |
| ON060775 | 201900877_L\|Uganda\|2018 | human | 2018 | Sembabule | C | C.2.2 |
| ON060796 | 201902737_L\|Uganda\|2018 | human | 2018 | Kampala | C | C.2.2 |
| ON060790 | 2019000825_L\|Uganda\|2018 | human | 2019 | Kampala | C | C.2.2 |
| ON060799 | 201902745_L\|Uganda\|2018 | human | 2018 | Mbarara | C | C.2.2 |
| MG972972 | 201707024 | human | 2017 | Mityana | C | C.2.2 |
| ON060778 | 201900879_L\|Uganda\|2018 | human | 2018 | Mubende | C | C.2.2 |
| ON060772 | 2019000825_L\|Uganda\|2019 | human | 2019 | Kampala | C | C.2.2 |
| ON060781 | 201900880_L\|Uganda\|2018 | human | 2018 | Kabale | C | C.2.2 |

**Supplementary Table 3**. Lineage assignment of complete medium segment genomic sequences.

| Accession | Strain | Host | Year | Location | Lineage | Subclade |
| --- | --- | --- | --- | --- | --- | --- |
| PQ436084 | CD532/Isingiro1-2023 | human | 2023 | Isingiro | C | C.2.2 |
| PQ436081 | CD300/Kabale1-2022 | human | 2022 | Kabale | C | C.2.2 |
| ON060829 | 2020001195_M\|Uganda | human | 2020 | Kampala | C | C.2.2 |
| PQ436082 | CD301/Rubanda1-2022 | human | 2022 | Rubanda | C | C.2.2 |
| ON060800 | 201902745_M\|Uganda\|2018 | human | 2018 | Mbarara | C | C.2.2 |
| ON060809 | 201902753_M\|Uganda\|2018 | human | 2018 | Mbarara | C | C.2.2 |
| MG972976 | 201707023 | human | 2017 | Kiboga | C | C.2.2 |
| ON060820 | 201902764_M\|Uganda\|2018 | human | 2018 | Kampala | C | C.2.2 |
| ON060817 | 201902763_M\|Uganda\|2018 | human | 2018 | Mubende | K | K.1.2 |
| ON060814 | 201902760_M\|Uganda\|2018 | human | 2018 | Ibanda | K | K.1.2 |
| ON060803 | 201902747_M\|Uganda\|2018 | human | 2018 | Isingiro | K | K.1.2 |
| DQ380191 | Entebbe | human | 1944 | Entebbe | K | K.1.1 |
| OP146108 | RSA/OBP/RVFVSmithburn/LAV | human | 1944 | Entebbe | K | K.1.1 |
| MG659817 | Lunyo | mosquito | 1955 | Lunyo | M |  |
| KU167026 | Lunyo | mosquito | 1955 | Lunyo | M |  |
| ON060791 | 201902734_M\|Uganda\|2018 | human | 2018 | Mbarara | C | C.2.2 |
| ON060773 | 2019000825_M\|Uganda\|2019 | human | 2019 | Kampala | C | C.2.2 |
| MG972973 | 201707024 | human | 2017 | Mityana | C | C.2.2 |
| ON060806 | 201902750_M\|Uganda\|2018 | human | 2018 | Mbarara | C | C.2.2 |
| ON060776 | 201900877_M\|Uganda\|2018 | human | 2018 | Sembabule | C | C.2.2 |
| ON060832 | 2020002574_M\|Uganda | human | 2020 | Kampala | C | C.2.2 |
| ON060788 | 201902733_M\|Uganda\|2018 | human | 2018 | Mbarara | C | C.2.2 |
| ON060779 | 201900879_M\|Uganda\|2018 | human | 2018 | Mubende | C | C.2.2 |
| ON060782 | 201900880_M\|Uganda\|2018 | human | 2018 | Kabale | C | C.2.2 |

**Supplementary Table 4**. Lineage assignment of complete small segment genomic sequences.

| Accession | Strain | Host | Year | Location | Lineage | Subclade |
| --- | --- | --- | --- | --- | --- | --- |
| ON060838 | 2021001358_S\|Uganda | human | 2020 | Kampala | C | C.2.2 |
| PQ436077 | CD300/Kabale1-2022 | human | 2022 | Kabale | C | C.2.2 |
| ON060795 | 201902736_S\|Uganda\|2018 | human | 2018 | Sheema | K | K.1.2 |
| ON060804 | 201902747_S\|Uganda\|2018 | human | 2018 | Isingiro | K | K.1.2 |
| ON060815 | 201902760_S\|Uganda\|2018 | human | 2018 | Ibanda | K | K.1.2 |
| ON060818 | 201902763_S\|Uganda\|2018 | human | 2018 | Mubende | K | K.1.2 |
| ON060786 | 201901671_S\|Uganda | human | 2019 | Yumbe | K | K.1.2 |
| EU312121 | LunyoUGA55 | mosquito | 1955 | Lunyo | M |  |
| MG659989 | Lunyo | mosquito | 1955 | Lunyo | M |  |
| KU167025 | Lunyo | mosquito | 1955 | Lunyo | M |  |
| DQ380156 | Entebbe | human | 1944 | Entebbe | K | K.1.1 |
| DQ380157 | Smithburn | human | 1944 | Entebbe | K | K.1.1 |
| OP146109 | RSA/OBP/RVFVSmithburn/LAV | human | 1944 | Entebbe | K | K.1.1 |
| ON060821 | 201902764_S\|Uganda\|2018 | human | 2018 | Kampala | C | C.2.2 |
| MG953426 | 201601502 | human | 2016 | Kabale | C | C.2.2 |
| MG953425 | 201601292 | human | 2016 | Kamuganguzi | C | C.2.2 |
| ON060812 | 201902754_S\|Uganda\|2018 | human | 2018 | Sembabule | C | C.2.2 |
| ON060774 | 2019000825_S\|Uganda\|2019 | human | 2019 | Kampala | C | C.2.2 |
| ON060798 | 201902737_S\|Uganda\|2018 | human | 2018 | Kampala | C | C.2.2 |
| ON060792 | 201902734_S\|Uganda\|2018 | human | 2018 | Mbarara | C | C.2.2 |
| MG972974 | 201707024 | human | 2017 | Mityana | C | C.2.2 |
| ON060801 | 201902745_S\|Uganda\|2018 | human | 2018 | Mbarara | C | C.2.2 |
| ON060830 | 2020001195_S\|Uganda | human | 2020 | Kampala | C | C.2.2 |
| PQ436078 | CD301/Rubanda1-2022 | human | 2022 | Rubanda | C | C.2.2 |
| PQ436079 | CD304/Rubanda2-2023 | human | 2023 | Rubanda | C | C.2.2 |
| ON060824 | 2020000148_S\|Uganda | human | 2020 | Kampala | C | C.2.2 |
| ON060833 | 2020002574_S\|Uganda | human | 2020 | Kampala | C | C.2.2 |
| ON060789 | 201902733_S\|Uganda\|2018 | human | 2018 | Mbarara | C | C.2.2 |
| ON060783 | 201900880_S\|Uganda\|2018 | human | 2018 | Kabale | C | C.2.2 |
| ON060810 | 201902753_S\|Uganda\|2018 | human | 2018 | Mbarara | C | C.2.2 |
| ON060807 | 201902750_S\|Uganda\|2018 | human | 2018 | Mbarara | C | C.2.2 |
| MG972977 | 201707023 | human | 2017 | Kiboga | C | C.2.2 |
| ON060777 | 201900877_S\|Uganda\|2018 | human | 2018 | Sembabule | C | C.2.2 |
| ON060780 | 201900879_S\|Uganda\|2018 | human | 2018 | Mubende | C | C.2.2 |
| ON060827 | 2020000159_S\|Uganda | human | 2020 | Kampala | C | C.2.2 |
